# PRANA: A Deep Learning Method for Adapting Polygenic Risk Scores to Diverse Ethnic Groups

**DOI:** 10.64898/2026.07.12.26357860

**Authors:** Hagai Levi, Qin Wang, Manjeet K. Bolla, Joe Dennis, Irene L. Andrulis, Natalia Antonenkova, Chun Hang Au, Annelie Augustinsson, Laura E. Beane Freeman, Sabine Behrens, Marina Bermisheva, Clara Bodelon, Natalia V. Bogdanova, Stig E. Bojesen, Hermann Brenner, Ian W. Brock, Thomas Brüning, Helen Byers, Nicola J. Camp, Jose E. Castelao, Ji-Yeob Choi, Wendy K. Chung, NBCS Collaborators, Sarah V. Colonna, Fergus J. Couch, Kamila Czene, Mary B. Daly, Peter Devilee, Thilo Dörk, A. Heather Eliassen, Mikael Eriksson, D. Gareth Evans, Peter A. Fasching, Kierstin Faw, Manuela Gago-Dominguez, Montserrat García-Closas, Christopher A. Haiman, Ute Hamann, Mikael Hartman, Vikki Ho, Peh Joo Ho, Maartje J. Hooning, Reiner Hoppe, Sacha J. Howell, ABCTB Investigators, SGBCC Investigators, Hidemi Ito, Motoki Iwasaki, Anna Jakubowska, Helena Jernström, Vijai Joseph, Rudolf Kaaks, Daehee Kang, Elza K. Khusnutdinova, Sanja Kiprijanovska, Stella Koutros, Vessela N. Kristensen, Allison W. Kurian, Ava Kwong, Diether Lambrechts, Nicole L. Larson, Loic Le Marchand, Shuai Li, Jingmei Li, Artitaya Lophatananon, Arto Mannermaa, Keitaro Matsuo, Whitney Maxwell, Kenneth Muir, Ines Nevelsteen, Nadia Obi, Katie M. O’Brien, Kenneth Ofit, Penelope D. Ottewell, Alpa V. Patel, Paolo Peterlongo, Dijana Plaseska-Karanfilska, Karolina Prajzendanc, Paolo Radice, Dhanya Ramachandran, Muhammad U. Rashid, Atocha Romero, Emmanouil Saloustros, Dale P. Sandler, Ben Schöttker, Jacques Simard, Melissa C. Southey, Jennifer Stone, Pierre-Emmanuel Sugier, Jack A. Taylor, Lauren R. Teras, Thérèse Truong, Clarice R. Weinberg, Siddhartha Yadav, Taiki Yamaji, Wei Zheng, Alison M. Dunning, Nasim Mavaddat, Douglas F. Easton, Kyriaki Michailidou, Ran Elkon, Ron Shamir

## Abstract

Polygenic risk scores (PRSs), which quantify inherited susceptibility to complex traits and diseases, have emerged as valuable tools for risk stratification and precision medicine. Despite their promise, PRS developed on European cohorts often demonstrate substantially reduced predictive accuracy in non-European populations, due to differences in genetic architecture. The disproportionate representation of European ancestry cohorts in genome-wide association studies (GWAS) leads to inequitable deployment of PRS technologies across diverse populations. Here, we introduce PRANA (Polygenic Risk Adaptation via Neural-network Architecture), a deep learning framework that adapts an existing PRS developed on one population to other ancestries. Unlike methods that require large-scale GWAS in the target population, PRANA leverages pre-trained PRS models derived from European cohorts and adapts them using modestly sized cohorts from the target population.

We evaluated PRANA on seven complex traits in South Asian, East Asian and Ashkenazi Jewish populations, as well as in selected smaller East Asian subpopulations where the scarcity of training data poses a particular challenge. PRANA mostly improved predictive performance of the baseline PRS models by 5%-20% in terms of effect size (β) and Nagelkerke’s R², and, in most cases, outperformed existing cross-ancestry multi-PRS approaches. These results highlight PRANA as a scalable and practical strategy to reduce disparities in genomic risk prediction and advance the equitable application of PRS in diverse populations.

## Introduction

Polygenic risk scores (PRSs) provide a quantitative measure of an individual’s inherited susceptibility to complex traits and diseases by aggregating the effects of numerous SNPs, typically numbering from several dozen to hundreds of thousands. Their increasing use in biomedicine and clinical research has underscored their potential utility in risk stratification, precision medicine, and early disease prediction (Khera et al., 2018; Lewis and Vassos, 2020).

A critical challenge in deploying PRS is the well-documented observation that PRSs trained on individuals of one ancestry perform worse in individuals of other populations, where performance declines with increased genetic distance between the two populations. At present, there is a substantial bias towards the European (EUR) population on which the majority of GWASs were performed, and thus, decreased PRS performance on non-European populations, including African (AFR), South Asian (SAS), and East Asian (EAS) ancestries (Martin *et al*., 2019; Duncan *et al*., 2019). This disparity, arising from differences in allele frequencies, linkage disequilibrium (LD) patterns, and genetic architecture, hinders the equitable deployment of PRS globally across populations.

To address this issue, a variety of methods have been proposed in recent years. Among them are (1) multi-PRS approaches, which aggregate scores derived from multiple populations (Márquez-Luna et al., 2017), (2) PRS-CSx, a Bayesian framework that leverages LD panels from different ancestries to jointly estimate SNP effects (Ruan *et al*., 2021), (3) PolyPred, which uses linear combinations of multiple ancestry-specific predictors, often incorporating functional annotations to improve portability (Weissbrod *et al*., 2022) and (4) transfer learning-based methods such as PRS-TL, which adapt effect sizes learned from one population to another using gradient descent (Lu et al., 2022). Each of these methods has demonstrated partial success, improving PRS performance in underrepresented populations to varying degrees. However, their effectiveness is frequently constrained by the limited sample sizes available for non-EUR GWASs, model complexity, and the intrinsic difficulty of capturing subtle ancestry-specific signals without overfitting.

Given the limitations of cross-ancestry prediction, recent efforts have begun exploring the potential of deep learning to enhance polygenic risk prediction. For example, PRS-Net (Li *et al*., 2025) attempts to incorporate global information from biological networks in graph neural networks (GNN), and uses attentive module for cross-ancestry prediction. But while deep learning has shown transformative success in fields such as computer vision, natural language processing, and drug discovery, in genomics, and in PRS modeling in particular, deep learning has not consistently outperformed classical statistical approaches (Schuran *et al*., 2025; Yuan *et al*., 2025). This is largely due to structural challenges in genomic data: datasets are often high-dimensional and sparse, signals are polygenic, and training cohorts are relatively small compared to other domains where deep learning thrives.

In this study, we introduce Polygenic Risk Adaptation via Neural-network Architecture (PRANA), a novel deep learning framework designed to adapt an existing PRS trained on one population to individuals from other populations by learning ancestry-specific transformations of the PRS signal. Rather than developing an entirely new model from raw genotype data, our method utilizes a pre-trained PRS model derived from a large-scale, typically European, GWAS, and uses a much smaller cohort, of just a few thousand individuals, from the target population to adapt the polygenic scores. We evaluated the performance of our method using PRS models generated by standard tools such as pruning and thresholding (P+T) (Purcell *et al*., 2007), Lassosum (Mak *et al*., 2017), or PRS-CSx (Ruan *et al*., 2021) on seven traits/diseases across four populations. In addition, we assessed the performance of PRANA within subpopulations, where data availability for the target subpopulations is limited.

## Methods

In the following section we describe the construction of PRS models, and the adaptation of these models to other populations. We call the large (typically EUR) population on which the initial PRS was developed the **discovery population**, and the population of a different ethnicity to which the PRS is to be adapted the **target population**.

### Datasets and GWAS summary statistics

#### UK Biobank

We ran GWASs for multiple binary phenotypes on the EUR population using UK Biobank (UKB) data (Bahcall, 2018), constructed PRS models for each phenotype, and adapted them to the target SAS population from the UKB. We started by performing QC using PLINK (Purcell *et al*., 2007; Chen *et al*., 2019), and kept only SNPs with MAF≥5%, HWE ≥ 1×10^-6^ and missing rate ≤10%. We kept only samples where less than 10% of the SNPs measured for the entire UKB cohort were missing. In addition, we filtered out ambiguous and duplicated alleles. 5,243,085 SNPs passed this process. The EUR GWASs utilized the entire EUR population from the UKB that passed our filtering (n=487,409). For SAS, we used 5000 individuals to perform GWASs. The rest of the SAS UKB cohort (n=4,416) was used for PRS model training and testing.

The phenotypes we analyzed using UKB were based on the following considerations: We extracted candidate diseases from the “non-cancer illness” field (code 20002), excluding Cholesterol and Hypertension, which are quantitative in nature. We filtered the diseases based on two criteria: (1) The estimated SNP-based heritability of the disease in the EUR GWAS we generated was above 2%, as measured by LDSC (Bulik-Sullivan *et al*., 2015) with default parameters. (2) The disease had > 500 cases in the UKB SAS population. Five diseases passed this filtering process: Type 2 Diabetes, Asthma, Hypothyroidism, Angina pectoris, and hay-fever. See Table S1 for the cohort statistics of each disease.

#### Schizophrenia - Ashkenazi Jewish (AJ) dataset

For the evaluation of Schizophrenia (SCZ) PRS in AJ, we used a EUR SCZ GWAS (Ripke *et al*., 2014) that excluded the AJ individuals. These individuals were used as the target set in our analysis (dbGaP access id: phs000448.v1.p1; 1044 cases, 2052 controls).

#### Breast Cancer Association Consortium (BCAC)

We extracted two discovery sets from BCAC (Michailidou *et al*., 2017): (1) EUR discovery set, comprising 72,899 BC cases and 59,436 controls, and (2) a small-scale EAS (Singaporean) discovery set, containing 3,566 BC cases and 3,343 controls. For each discovery set, we applied the same QC as for the UKB data. 4,954,903 and 5,649,407 SNPs passed this process, for EUR and EAS, respectively. The remaining EAS genotypes (n=13,241; 7607 cases, 5634 controls) were used as the target set in our analysis.

### Single-ethnic PRS methods

We used two methods for constructing PRS models using GWAS data from a single population: (1) Pruning and thresholding (P+T), using PLINK (Purcell *et al*., 2007), and (2) Lassosum (Mak *et al*., 2017). For each method, we chose a set of hyperparameters that yielded PRS models with < 500 SNPs. In the UKB, the Angina and Hay-Fever GWASs had weaker signals compared to the other three diseases (<75 SNPs with p-value < 1×10^-08^, and <500 SNPs with p-value < 1×10^-04^). To avoid very sparse models, we chose two sets of hyper-parameters – one for the stronger GWASs (Type 2 Diabetes, Asthma, Hypothyroidism) and another for the weaker ones (Angina, Hay-Fever). Details on the selected models and hyperparameters are provided in Table S2.

### Multi-PRS methods

PRANA was evaluated against several multi-PRS methods on the BCAC EAS population: (1) multi-pruning and thresholding (P+T), using PLINK (Purcell *et al*., 2007), (2) multi-Lassosum (Mak *et al*., 2017), and (3) PRS-CSx (Ruan *et al*., 2021). These methods receive a large-scale EUR GWAS as input, and a small-scale EAS GWAS and construct PRS for each of them. PRS-CSx was applied using the two GWASs jointly in the same execution. Each method yielded two sets of SNP weights - one from the EUR GWAS (EUR PRS), and one from the EAS GWAS (EAS PRS). In all three methods the combined PRS was obtained by fitting linear regression on the two sets of weights.

We constructed multiple EUR- and EAS-PRS models using different configurations of hyperparameters (Table S3), and applied nested cross-validation to identify best-performing model configurations, selecting one optimized model per ancestry.

### Nested cross-validation scheme

We applied a modified version of the (*n* + 1) ∗ *n* nested cross validation (CV) scheme described in (Levi *et al*., 2023). Briefly, each target set cohort was split into *n* + 1 subsets. One subset was held out as an external test set, while the remaining n subsets were used for an internal *n*-fold CV, where PRS models were trained on *n* − 1 folds and evaluated on the remaining validation fold. After iterating over the *n* combinations of training and validation folds, the best hyper-parameter set was selected based on average performance on the validation sets. The final PRS model was retrained on all *n* folds with the best hyperparameter set and evaluated on the test set. This entire process was repeated *n* + 1 times, each time designating a different subset as the test set, and the performance on the test set was averaged across all iterations.

The multi-PRS methods use GWAS summary statistics derived from a subset of the target population (target population discovery set), reserving the remaining genotypes for hyperparameter tuning. In contrast, PRANA does not use GWAS summary statistics, and therefore, we included all the genotypes of the target population discovery set directly as PRANA’s training set for both the UKB and the BCAC datasets.

We utilized a 4X3 nested CV strategy for the UKB and the SCZ-AJ datasets. For the BCAC cohort, where the effective sample size was larger, we applied a 6X5 nested CV scheme.

### Evaluation metrics

We used four criteria to evaluate the performance of PRS models: Association coefficient (Beta), Nagekerke’s R^2^, Area Under the Receiver Operating Curve (AUROC), and Area Under Precision Recall Curve (AUPRC). To calculate Beta and Nagelkerke’s R^2^, we trained a logistic regression model that predicts the disease status from the PRS and top five principal components (PCs) obtained by PCA as performed previously (Levi *et al*., 2024).

### The PRANA architecture

The outline of PRANA is described in Figure 1. Given a PRS model comprising *n* SNPs, PRANA one-hot encodes the genotypes as a 3*xn* bit matrix. A SNP genotype homozygous to the reference is as [1,0,0], heterozygous encoded as [0,1,0], and homozygous to the alternative is encoded as [0,0,1]. Missing genotypes are encoded [0,0,0]. Each encoded SNP is passed into a 3-to-1 linear layer, with initial edge weights depending on the minor allele frequency (MAF) of the SNP, as follows: we set the weight associated with each dosage to *dosage* − 2 ⋅ *MAF*_train_ where dosage=0/1/2, and the intercept to 2 *MAF*_train_, where *MAF*_train_ is the MAF calculated on the training set data.

**Figure 1.**
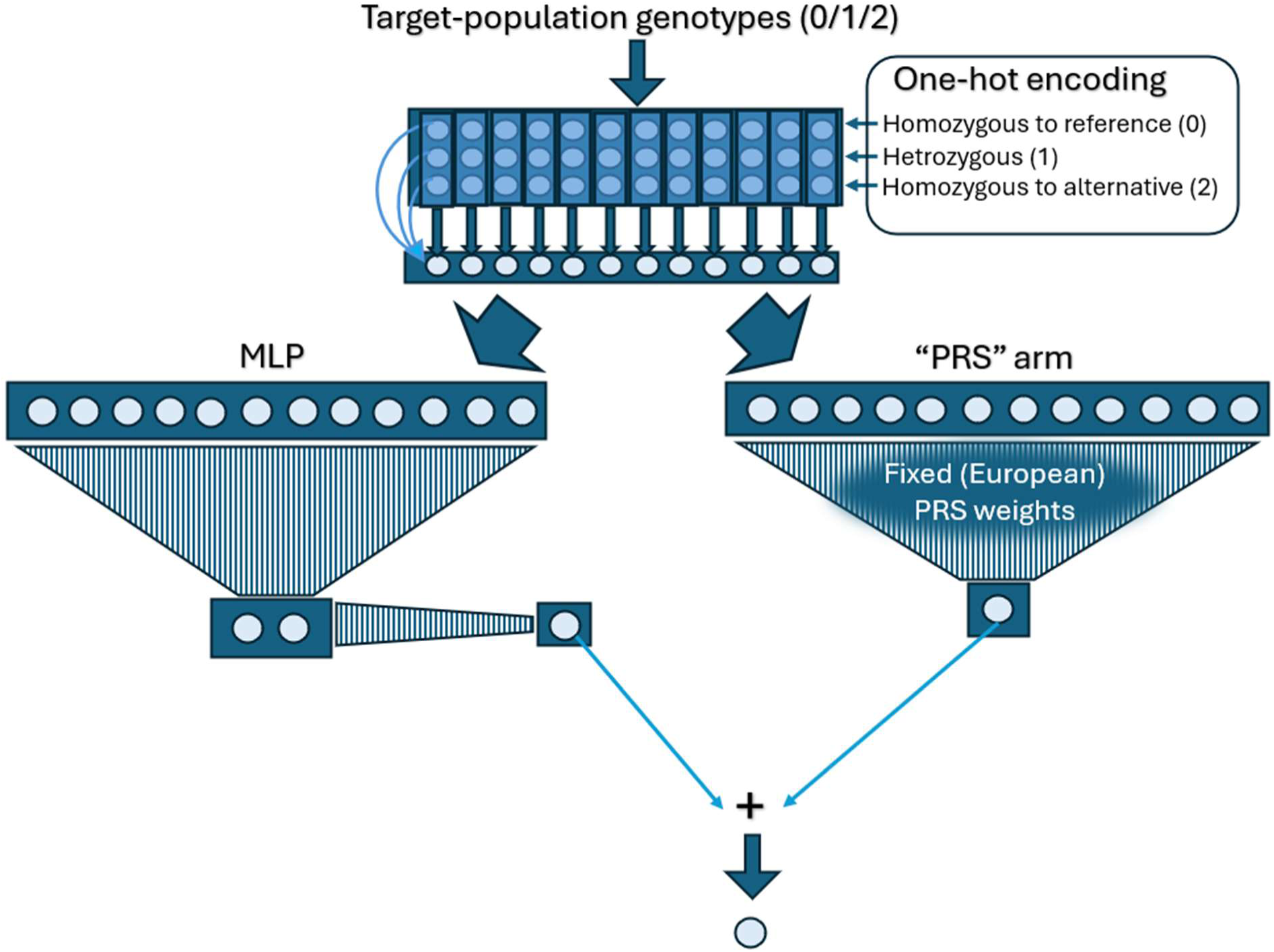
Outline of PRANA. The method one-hot encodes genotypes as input, weighting them by homozygous/heterozygous status to produce a dosage value per SNP and weighted by MAF. This encoding accounts for missing data and potential non-additive effects (see Methods). The encoded values are then passed through two parallel arms: (1) a fixed ‘PRS’ arm with weights initialized from an external pre-computed PRS model and kept frozen, and (2) a 2-layer MLP forming the learnable adapter arm. The values output by each arm are summed to generate the final predicted risk.

The encoded values are then passed through two parallel arms: (1) a linear n-to-1 perceptron called ‘PRS’, where the weights are frozen and are set according to the original base PRS model; and (2) a learnable fully connected *parallel adapter arm*. This arm is a multi-layer perceptron (MLP) with two layers: n-to-2, and 2-to-1. To ensure that the weights in the first layer in the adapter arm are scaled proportionally to those of the PRS arm, we used a modification of the weight initialization proposed by (Glorot and Bengio, 2010; He *et al*., 2015). Specifically, the weights of the first layer in the adapter arm were drawn from 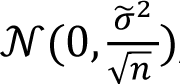, where 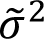 is the variance of the original PRS weights and *n* is the number of SNPs. No activation function was applied after any of the layers. Each arm outputs a single value, and the sum of the two values is the final predicted risk score.

### Selecting the number of epochs

Using the nested CV process, we select the optimal number of training epochs for PRANA from the set {0, 100, 200, 300}. For the larger BCAC cohort, since a larger number of folds allows more robust exploration of hyperparameter values, we used the set of values {0, 50, 100, 150, 200, 250, 300}.

### Loss function

We constructed a customized loss function inspired by Nagelkerke’s R^2^, which uses the log likelihoods of the logistic regression model trained to predict a trait from the risk scores:

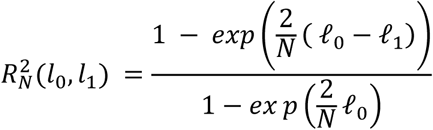

Where ℓ_1_ and ℓ_O_ are the log likelihoods of the tested and the null models, respectively, and N is the size of the batch for which we compute the loss.

For each batch, we approximated a logistic regression model by training another single-layer fully connected network, which we called LogiNet. It is fed with the predicted risk score and the top 5 PCs. The layer outputs a single value, which is input into a sigmoid activation function. LogiNet is trained for 100 epochs, with binary cross entropy (BCE) loss, Adam optimizer, and learning rate of 0.01. After the training, we compute LogiNet’s log likelihood (−*BCE*; denoted ℓ̂_1_), which is used in our loss function to backpropagate to the weights of PRANA. Next, ℓ̂_1_ is plugged in to compute an approximation of Nagelkerke’s 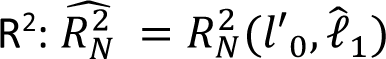, where ℓ′_O_ is computed from a logistic regression model fitted by the *statsmodels* package in Python with the same features. The final loss value is 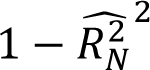. The squaring of 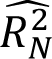 was performed to mitigate potential overfitting arising from the amplification of small estimation errors in 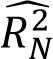. This loss function ultimately backpropagates gradients to update the PRANA weights through ℓ̂_1_, as ℓ̂_1_ is computed using the risk scores generated by PRANA.

For the UKB and the BCAC datasets, PCs were computed using independent ethnically heterogenous genotypes from the UKB and BCAC, respectively. For the SCZ-AJ datasets, where such independent data was not available, we used only the batch data to compute the PCs for the computation of that batch to avoid data leakage.

## Results

We developed PRANA (Figure 1 and Methods), a deep-learning method to improve risk prediction for binary outcomes in underrepresented target populations by adapting existing PRS models constructed from a larger, ancestrally distinct population. Starting from a PRS base model trained on a large-scale discovery cohort, typically available for EUR population, PRANA fixes a “PRS arm” according to the weights of the base model. The transfer is performed through a *parallel adapter arm*, inspired by (Houlsby *et al*., 2019; Rebuffi *et al*., 2018). The adapter arm comprises learnable weights that are fine-tuned using genotypes of a modest number of individuals in the target population—without requiring additional GWAS summary statistics. Importantly, PRANA does not add or remove SNPs from the original base model, which ensures that (1) the required SNPs for the target population are known a priori, and (2) model parsimony is maintained by avoiding an expansion of the set of input SNPs. This framework aims to narrow the gap between well-powered PRS models, typically derived from European cohorts, and the lack of such predictive models in diverse populations with limited cohort sizes.

To test the ability of PRANA to adapt existing PRS model to a different target population, we measured the relative increase in prediction performance when adapting base EUR PRS to non-EUR populations. We began by testing PRANA in SAS population from the UKB (Bahcall, 2018) across five binary traits: Type 2 Diabetes, Asthma, Hypothyroidism, Angina, and Hay fever (See Table S1 for cohort sizes). For each trait, we constructed baseline PRSs from EUR GWAS summary statistics using two popular PRS construction methods: (1) pruning and thresholding (P+T) (Purcell *et al*., 2007), and (2) Lassosum (Mak et al., 2017). Then, we used PRANA to adapt each PRS to the target SAS population. The adapted models were trained and evaluated using a nested cross validation scheme (see Methods), with the following metrics: (1) Beta (PRS association coefficient), (2) Nagelkerke’s R^2^, (3) Area Under Receiver Operating Curve (AUROC), and (4) Area Under Precision Recall Curve (AUPRC).

Figure 2 shows the relative improvement achieved by PRANA adaptation of the original EUR-based PRS model to the SAS population. PRANA improved performance as measured by all the four indexes: compared to the EUR base PRS model, Beta, Nagelkerke’s R^2^ AUROC and AUPRC were improved, respectively, in 9, 8, 8, and 6 out of the 10 tested PRS method-trait combinations (Figure 2).

**Figure 2.**
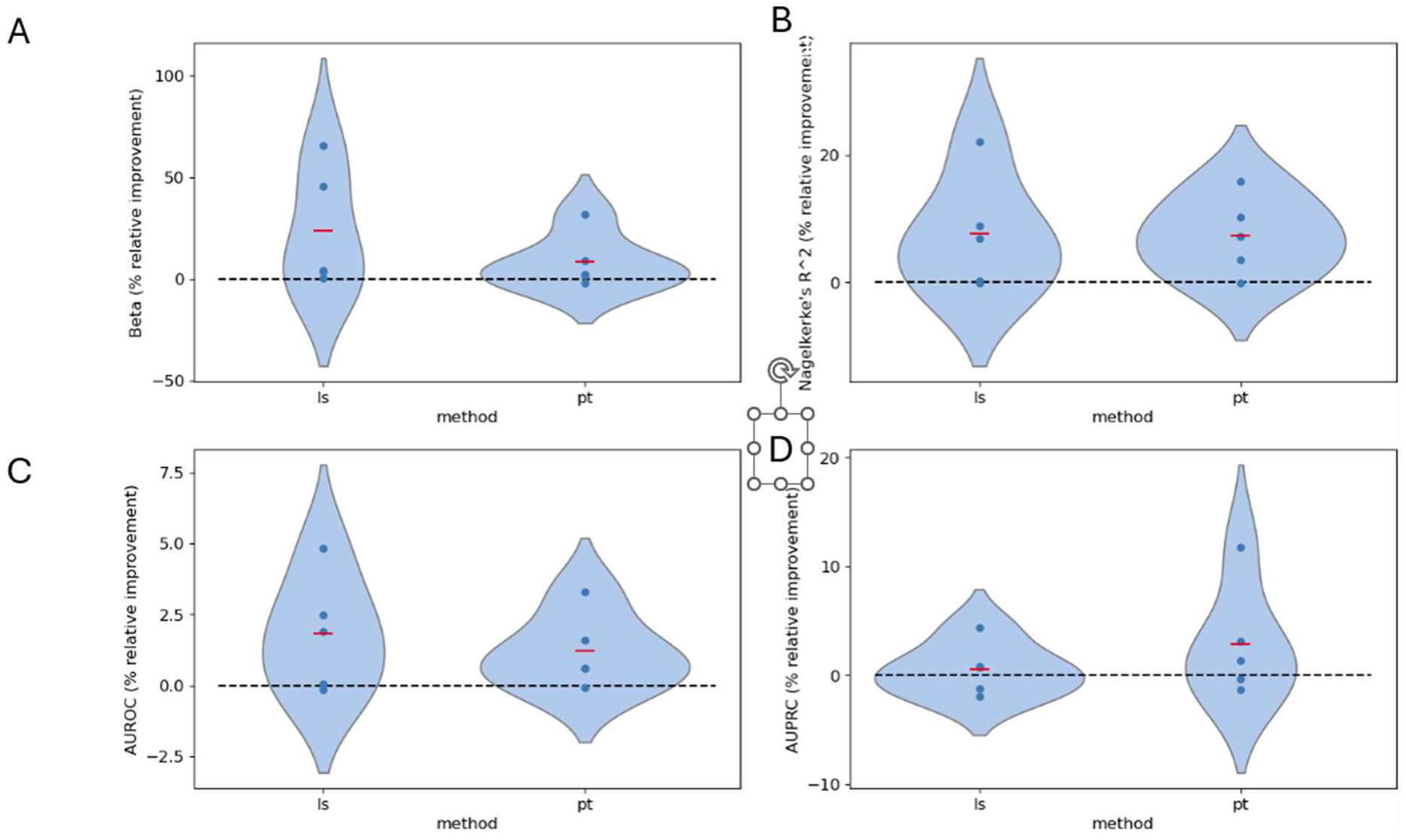
Relative improvement of PRANA compared to the original PRS models for five binary traits in SAS population from the UKB. P+T (pt), and lassosum (ls). Red dashes are the means.

Next, we constructed SAS-based PRS models for these five traits using P+T (Purcell *et al*., 2007) and lassosum (Mak *et al*., 2017). For this aim, we ran GWAS analysis using 5,000 SAS individuals from the UKB (Table S1) and constructed PRS for SAS. We then compared the performance of the EUR-based PRS model adapted by PRANA against (1) the original EUR PRS model, (2) the SAS PRS model, and (3) a combined PRS model that incorporates the risk scores computed by EUR and SAS PRSs as two features in a linear regression. These models were tested using nested CV on the remaining SAS cohort (n= 4416. See Methods). Table 1 summarizes the results. In 8 out of 10 cases, PRANA achieved the best performance in Beta, Nagelkerke’s R^2^ and AUROC. Notably, both the SAS and the combined PRS models rely on a substantially larger set of SNPs (Table S2) than the PRANA-adapted model, underscoring the ability of PRANA to improve prediction even with a more parsimonious SNP set.

**Table 1.** Performance comparison of the EUR-based PRS model adapted by PRANA against (1) the original EUR PRS model, (2) the SAS PRS model, and (3) a combined PRS model that incorporates the risk scores computed by EUR and SAS PRSs.

| Base method | Trait | Method | Beta | Nagelkerke's R<sup>2</sup> | AUROC | AUPRC |
| --- | --- | --- | --- | --- | --- | --- |
| Lassosum | angna | PRANA | **0.255±0.031** | **0.031±0.004** | **0.569±0.007** | 0.064±0.005 |
|  | angna | ls-combined | 0.219±0.008 | 0.03±0.006 | 0.567±0.012 | 0.078±0.008 |
|  | angna | ls-eur | 0.162±0.023 | 0.026±0.005 | 0.543±0.009 | 0.066±0.006 |
|  | angna | ls-sas | 0.161±0.005 | 0.026±0.006 | 0.546±0.011 | **0.082±0.01** |
|  | ast | PRANA | **0.186±0.06** | **0.021±0.006** | **0.556±0.017** | **0.149±0.014** |
|  | ast | ls-combined | 0.166±0.073 | 0.02±0.006 | 0.544±0.022 | 0.145±0.014 |
|  | ast | ls-eur | 0.163±0.06 | 0.019±0.006 | 0.543±0.017 | 0.143±0.013 |
|  | ast | ls-sas | 0.057±0.052 | 0.014±0.006 | 0.516±0.01 | 0.129±0.008 |
|  | hfvr | PRANA | **0.131±0.051** | 0.025±0.008 | **0.536±0.012** | 0.063±0.005 |
|  | hfvr | ls-combined | 0.088±0.038 | 0.023±0.008 | 0.534±0.008 | 0.063±0.0 |
|  | hfvr | ls-eur | **0.131±0.051** | 0.025±0.008 | **0.536±0.012** | 0.064±0.002 |
|  | hfvr | ls-sas | 0.059±0.081 | **0.026±0.008** | 0.528±0.026 | **0.07±0.008** |
|  | hyty | PRANA | **0.409±0.067** | **0.048±0.008** | **0.611±0.016** | **0.1±0.007** |
|  | hyty | ls-combined | 0.403±0.064 | **0.048±0.008** | **0.611±0.017** | **0.1±0.009** |
|  | hyty | ls-eur | 0.39±0.046 | 0.045±0.006 | 0.599±0.014 | **0.1±0.007** |
|  | hyty | ls-sas | 0.152±0.05 | 0.026±0.006 | 0.544±0.018 | 0.08±0.004 |
|  | t2d | PRANA | 0.229±0.043 | 0.033±0.007 | 0.573±0.016 | 0.189±0.005 |
|  | t2d | ls-combined | **0.251±0.054** | **0.036±0.008** | **0.575±0.019** | **0.192±0.006** |
|  | t2d | ls-eur | 0.228±0.043 | 0.033±0.007 | 0.573±0.016 | 0.188±0.005 |
|  | t2d | ls-sas | 0.115±0.042 | 0.024±0.007 | 0.534±0.011 | 0.168±0.006 |
| P+T | angna | PRANA | **0.263±0.015** | **0.032±0.007** | **0.579±0.01** | **0.074±0.008** |
|  | angna | pt-combined | 0.24±0.029 | 0.03±0.006 | 0.574±0.007 | 0.07±0.004 |
|  | angna | pt-eur | 0.201±0.012 | 0.028±0.006 | 0.56±0.005 | 0.066±0.005 |
|  | angna | pt-sas | 0.126±0.042 | 0.024±0.006 | 0.547±0.007 | 0.067±0.005 |
|  | ast | PRANA | **0.22±0.057** | **0.023±0.006** | **0.557±0.022** | **0.152±0.011** |
|  | ast | pt-combined | 0.174±0.053 | 0.019±0.006 | 0.541±0.02 | 0.145±0.009 |
|  | ast | pt-eur | 0.212±0.052 | 0.022±0.006 | 0.553±0.02 | 0.15±0.01 |
|  | ast | pt-sas | -0.159 | 0.015±0.008 | 0.472±0.016 | 0.117±0.007 |
|  | hfvr | PRANA | 0.097±0.053 | 0.024±0.006 | 0.521±0.018 | 0.069±0.004 |
|  | hfvr | pt-combined | 0.13±0.052 | 0.025±0.007 | 0.519±0.018 | **0.075±0.007** |
|  | hfvr | pt-eur | 0.104±0.059 | 0.024±0.006 | 0.522±0.019 | 0.069±0.004 |
|  | hfvr | pt-sas | **0.146±0.063** | **0.026±0.007** | **0.524±0.017** | 0.07±0.001 |
|  | hyty | PRANA | **0.454±0.046** | **0.052±0.009** | **0.625±0.01** | **0.097±0.006** |
|  | hyty | pt-combined | 0.432±0.06 | 0.049±0.011 | 0.617±0.015 | 0.092±0.004 |
|  | hyty | pt-eur | 0.425±0.057 | 0.048±0.011 | 0.615±0.014 | 0.094±0.005 |
|  | hyty | pt-sas | 0.07±0.034 | 0.022±0.006 | 0.526±0.012 | 0.071±0.003 |
|  | t2d | PRANA | **0.165±0.067** | **0.029±0.007** | **0.557±0.016** | 0.17±0.005 |
|  | t2d | pt-combined | 0.16±0.051 | 0.027±0.006 | 0.551±0.016 | **0.176±0.007** |
|  | t2d | pt-eur | 0.147±0.046 | 0.026±0.007 | 0.554±0.012 | 0.172±0.004 |
|  | t2d | pt-sas | 0.087±0.042 | 0.023±0.004 | 0.524±0.013 | 0.165±0.008 |

We next evaluated PRANA on additional datasets and ethnicities. First, we evaluated it on Schizophrenia (SCZ) in a cohort of Ashkenazi Jews (AJ) (Lencz *et al*., 2013). As before, we generated PRS from EUR SCZ GWAS (Ripke *et al*., 2014) using P+T and Lassosum (Methods). Consistent with the previous results, PRANA achieved higher predictive performance than the original EUR base model, reaching >30% relative improvement in Beta (Figure 3).

**Figure 3.**
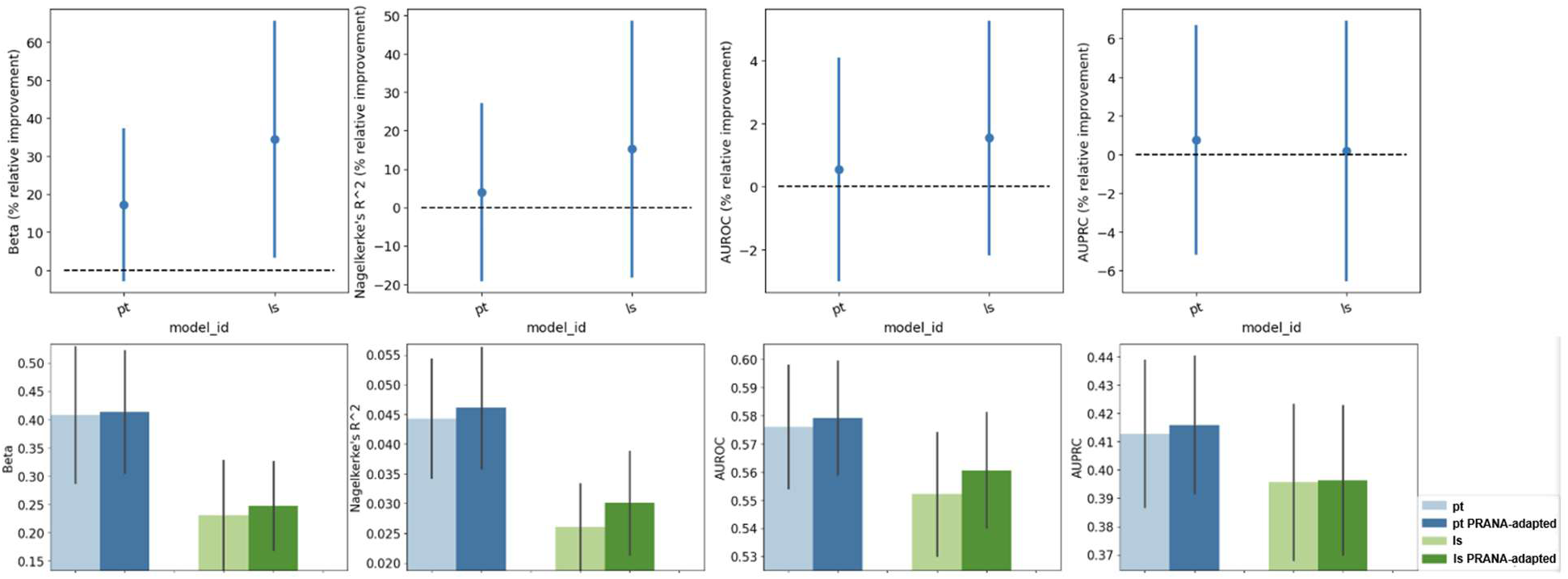
Predictive performance of Schizophrenia PRS model in Ashkenazi Jews. Top: relative improvement of PRANA vs. the baseline PRS method. Bottom: absolute values for the baseline PRS methods and the PRANA-adapted models. Error bars are SEM across folds. pt: Pruning and thresholding, ls: Lassosum.

Next, we evaluated our method for the breast cancer (BC) using the EAS cohort from the BCAC as the target population. This relatively large non-EUR cohort includes 11,173 BC cases and 8,977 controls of EAS ancestry. In particular, the high number of cases, enabled us to conduct nested CV with a greater number of folds, allowing a more extensive hyperparameter search (see Methods). Using PRANA, we generated an EAS-adapted version of the established 313-SNPs BC PRS (BC PRS_313_) constructed by Mavaddat et al. (Mavaddat et al., 2019). In this analysis too, the PRS model adapted by PRANA markedly improved the performance of the original BC PRS_313_ model on EAS women, obtaining 6.22 ± 3.58, 12.35 ± 7.61, 1.17 ± 0.71, and 1.26 ± 0.7 percent improvement in Beta, Nagelkerke’s R^2^, AUROC, and AUPRC, respectively (mean ± SEM). These results are slightly higher than those obtained using the original hyperparameter values (Table S4).

Using the BCAC dataset, we were also able to compare the BC PRS_313_ model adapted by PRANA for EAS women to multi-PRS models constructed by three popular multi-PRS methods: multi-PT, multi-lassosum, and PRS-CSx (Ruan *et al*., 2021) (Methods). For the construction of PRS models by these methods, we used BC GWASs we generated for the EUR and the EAS BCAC cohorts as input (see Methods). Notably, even though PRANA relies only on a limited set of SNPs (n=313), it outperformed the other methods in terms of Nagelkerke’s R2, AUROC, and AUPRC (Figure 4). The Beta value obtained by PRANA was second only to the model constructed by PRS-CSx, which included hundreds of thousands of SNPs. These findings are particularly important, as they show that when a high-quality small EUR-PRS model is already available, PRANA can be used to generate a lightweight PRS model that outperforms or is comparable to other multi-PRS models, which typically use numerous SNPs.

**Figure 4.**
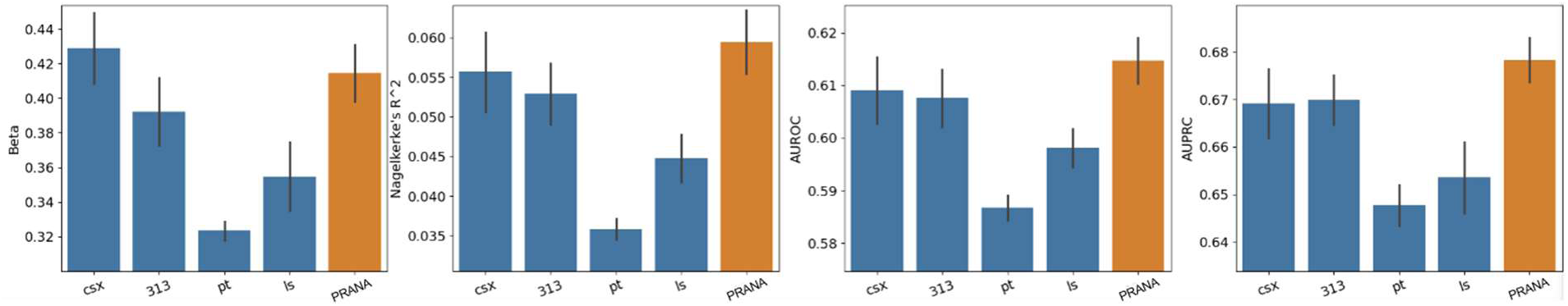
Predictive performance of breast cancer PRS in EAS population from the BCAC dataset. PRS methods: PRS-CSx (csx), Mavaddat’s 313-PRS (313), multi-P+T (pt), multi-lassosum (ls) and PRANA. Error bars are SEM. Epoch jumps=50.

We then evaluated PRANA performance across different EAS subpopulations by stratifying the original BCAC EAS cohort by country of origin (see Table S5 for subpopulation sizes). For each country, the PRANA model was trained on samples excluding that country and then tested on the samples from the held-out country. In this analysis too, the EUR BC PRS_313_ was used as the base model. Figure 5 shows performance as a function of the number of training epochs. For most countries the largest improvement was obtained between 20 and 40 epochs. These results highlight the ability of PRANA to utilize the information from the super-population, even when data for the target subpopulation is limited.

**Figure 5.**
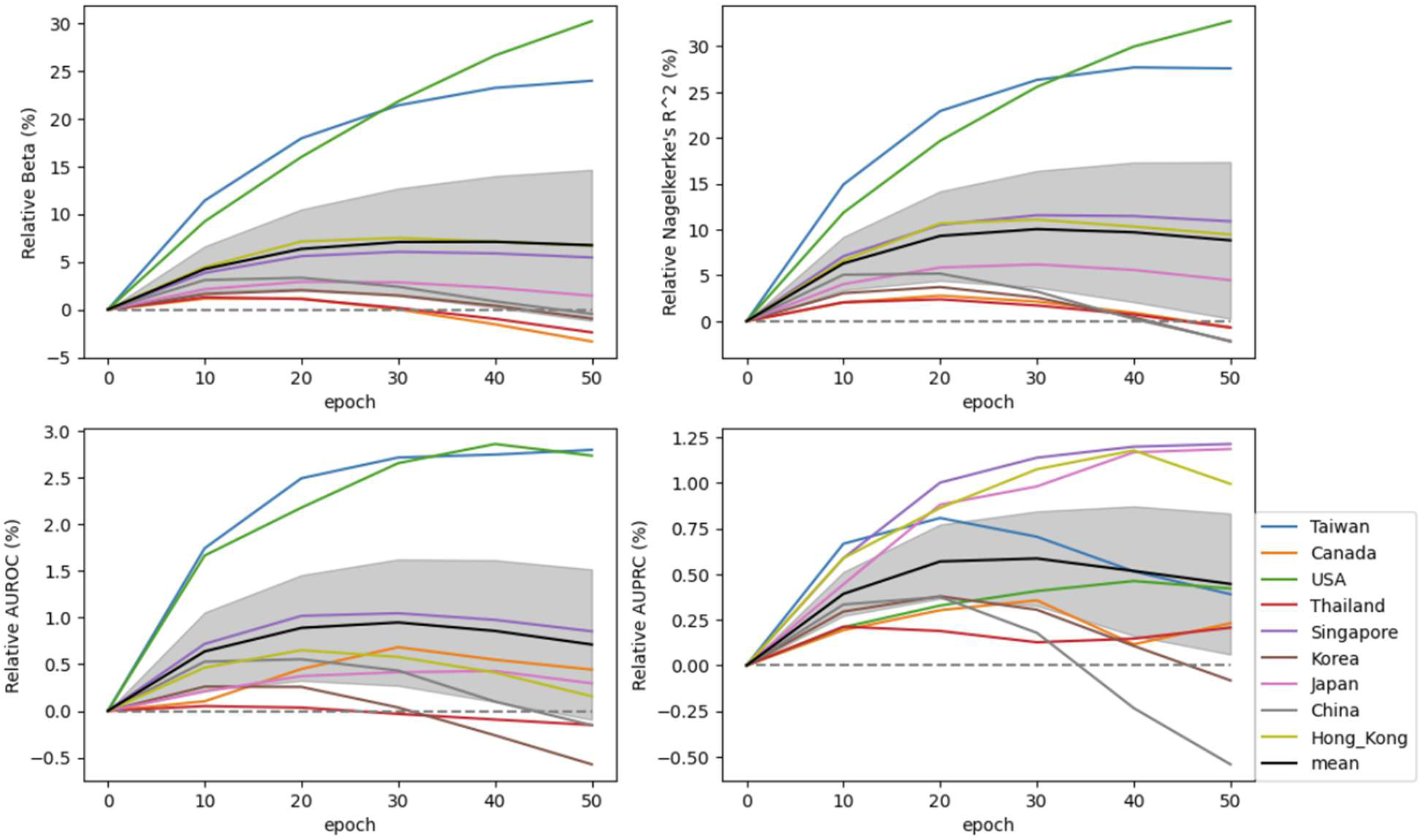
Predictive performance of PRANA across training epochs on breast cancer in different East Asian subgroups, stratified by country. The USA and Canada groups consist of EAS immigrants. The black line is the mean across groups. The shaded area is the confidence interval of the mean. A maximum of 50 epochs was chosen in applying the models based on the most frequent best number of epochs chosen in the nested CV.

Taken together, our results demonstrate that PRANA improves prediction in underrepresented populations by recalibrating lightweight EUR PRSs. This improvement persists even within subpopulations where available data is limited. We also show the ability of PRANA to process PRS models that were developed in the general population and adapt them for risk prediction in target groups who carry rare mutations. Moreover, in most cases PRANA’s performance matched or exceeded multi-PRS approaches, which construct high-dimensional models that require extensive GWAS summary data.

## Discussion

Polygenic risk scores have shown great promise with clear clinical potential. However, while they achieve strong predictive performance when trained on large cohorts - currently available mainly for European ancestry populations - their accuracy drops in non-European target populations, with the decline closely tied to the genetic distance from the discovery population.

In this study, we introduced PRANA, a deep learning method designed to adapt existing PRS models to new target populations. PRANA requires only a few thousand genotypes for the adaptation, an advantage given that GWAS derived from such small sample sizes are typically very noisy. Furthermore, PRANA preserves the original PRS feature set, enabling broader applicability across different contexts and ensuring that the adapted models remain lightweight. By utilizing a highly parsimonious SNP set, PRANA-adapted models lower computational overhead and simplify data-handling requirements, making them substantially easier to implement within real-world clinical workflows and public healthcare systems where resource efficiency is paramount.

We evaluated PRANA across four datasets, each representing a different target population, and observed consistent improvements in predictive performance. Additionally, our results indicate that PRANA enhances prediction accuracy even when only genotypes from a closely related subpopulation are available, underscoring its utility in real-world scenarios with limited data. Usually, learning plateaued on the held-out subpopulation after 20-30 epochs. Interestingly, the only population in which PRANA continued to improve after 50 epochs was the EAS immigrants from the USA (Figure 5). A possible explanation is the distance between the training and target populations: Previous studies have shown that including additional samples in the training process improves performance, particularly when these samples are ethnically matched to the target set (Ruan *et al*., 2021). This likely reflects the higher relevance of the training data to this specific group; as the training set contains a high density of samples matching the ancestral background of US-based EAS individuals, the model can extract more complex features before plateauing.

In addition, we compared PRANA with popular multi-PRS models. Despite relying on a much smaller set of SNPs, PRANA achieved superior performance in most cases and across multiple metrics. These results highlight the advantages of adaptation of an extant PRS model over constructing a new model for the target population from small GWASs.

One limitation of PRANA is its runtime. While training remains efficient for a few hundred SNPs, the computational cost increases when the set grows to thousands of SNPs (Figure S1). As a result, PRANA is best suited for adapting small- to medium-sized PRS models, and less so for those containing tens of thousands of SNPs.

Beyond pure ancestry, an active field of research also explores how to combine PRS models with non-genetic variables, such as the shifting lifestyle factors in different ethnic groups. Excitingly, PRANA’s deep learning architecture is highly flexible and could potentially be extended in future iterations to explicitly capture these complex gene-environment interactions to further refine risk prediction.

In summary, our results demonstrate that PRANA effectively improves the predictive performance of binary trait PRSs across diverse and underrepresented ancestral groups.

## Funding

This study was supported in part by grants from the Israeli Science Foundation (No. 3165/19, within the Israel Precision Medicine Partnership program, and No. 2206/22, to RS), from the Tel Aviv University Center for AI and Data Science (TAD) to RE and RS, by a joint program grant from the Cancer Biology Research Center, Djerassi Oncology Center, Edmond J. Safra Center for Bioinformatics and TAD to RE, and by the Koret-UC Berkeley-Tel Aviv University Initiative in Computational Biology and Bioinformatics to RE and RS. HL was supported in part by a fellowship from the Edmond J. Safra Center for Bioinformatics at Tel Aviv University.

## Data Availability

The source code of PRANA is available at: https://github.com/hag007/PRANA

The Breast Cancer Association Consortium (BCAC) data are available upon request from Cambridge University (see the BCAC website: https://bcac.ccge.medschl.cam.ac.uk/bcacdata/).

AJ SCZ genotypes were derived by permission from dbgap (https://www.ncbi.nlm.nih.gov/gap/) using access ids phs000021.v3 phs000448.v1.p1

The UK biobank data were used by permission from https://www.ukbiobank.ac.uk/

## BCAC Funding and Acknowledgments

### Funding

This work was supported by Cancer Research UK grant: PPRPGM-Nov20\100002 and by core funding from the NIHR Cambridge Biomedical Research Centre (NIHR203312) [*]. *The views expressed are those of the author(s) and not necessarily those of the NIHR or the Department of Health and Social Care. Additional funding for BCAC is provided by the Confluence project which is funded with intramural funds from the National Cancer Institute Intramural Research Program, National Institutes of Health, the European Union’s Horizon 2020 Research and Innovation Programme (grant numbers 634935 and 633784 for BRIDGES and B-CAST respectively), and the PERSPECTIVE I&I project, funded by the Government of Canada through Genome Canada and the Canadian Institutes of Health Research, the Ministère de l’Économie et de l’Innovation du Québec through Genome Québec, the Quebec Breast Cancer Foundation. The EU Horizon 2020 Research and Innovation Programme funding source had no role in study design, data collection, data analysis, data interpretation or writing of the report.

Genotyping of the OncoArray was funded by the NIH Grant U19 CA148065, and Cancer Research UK Grant C1287/A16563 and the PERSPECTIVE project supported by the Government of Canada through Genome Canada and the Canadian Institutes of Health Research (grant GPH-129344) and, the Ministère de l’Économie, Science et Innovation du Québec through Genome Québec and the PSRSIIRI-701 grant, and the Quebec Breast Cancer Foundation. Funding for iCOGS came from: the European Community’s Seventh Framework Programme under grant agreement n° 223175 (HEALTH-F2-2009-223175) (COGS), Cancer Research UK (C1287/A10118, C1287/A10710, C12292/A11174, C1281/A12014, C5047/A8384, C5047/A15007, C5047/A10692, C8197/A16565), the National Institutes of Health (CA128978) and Post-Cancer GWAS initiative (1U19 CA148537, 1U19 CA148065 and 1U19 CA148112 - the GAME-ON initiative), the Department of Defence (W81XWH-10-1-0341), the Canadian Institutes of Health Research (CIHR) for the CIHR Team in Familial Risks of Breast Cancer, and Komen Foundation for the Cure, the Breast Cancer Research Foundation, and the Ovarian Cancer Research Fund.

The Australian Breast Cancer Family Study (ABCFS) was supported by grant UM1 CA164920 from the National Cancer Institute (USA). The content of this manuscript does not necessarily reflect the views or policies of the National Cancer Institute or any of the collaborating centers in the Breast Cancer Family Registry (BCFR), nor does mention of trade names, commercial products, or organizations imply endorsement by the USA Government or the BCFR. The ABCFS was also supported by the National Health and Medical Research Council of Australia, the New South Wales Cancer Council, the Victorian Health Promotion Foundation (Australia) and the Victorian Breast Cancer Research Consortium. M.C.S. is a NHMRC Senior Research Fellow. The ABCS study was supported by the Dutch Cancer Society [grants NKI 2007-3839; 2009 4363] and an institutional grant of the Dutch Cancer Society and of the Dutch Ministry of Health, Welfare and Sport. The Australian Breast Cancer Tissue Bank (ABCTB) was supported by the National Health and Medical Research Council of Australia, The Cancer Institute NSW and the National Breast Cancer Foundation. The ACP study is funded by the Breast Cancer Research Trust, UK. KM and AL are supported by the NIHR Manchester Biomedical Research Centre, the Allan Turing Institute under the EPSRC grant EP/N510129/1. The AHS study is supported by the intramural research program of the National Institutes of Health, the National Cancer Institute (grant number Z01-CP010119), and the National Institute of Environmental Health Sciences (grant number Z01-ES049030). The work of the BBCC was partly funded by ELAN-Fond of the University Hospital of Erlangen. The BBCS is funded by Cancer Research UK and Breast Cancer Now and acknowledges NHS funding to the NIHR Biomedical Research Centre, and the National Cancer Research Network (NCRN). The BCEES was funded by the National Health and Medical Research Council, Australia and the Cancer Council Western Australia and acknowledges funding from the National Breast Cancer Foundation (JS). For the BCFR-NY, BCFR-PA, BCFR-UT this work was supported by grant UM1 CA164920 from the National Cancer Institute. The content of this manuscript does not necessarily reflect the views or policies of the National Cancer Institute or any of the collaborating centers in the Breast Cancer Family Registry (BCFR), nor does mention of trade names, commercial products, or organizations imply endorsement by the US Government or the BCFR. . The BREast Oncology GAlician Network (BREOGAN) is funded by Acción Estratégica de Salud del Instituto de Salud Carlos III FIS PI12/02125/Cofinanciado and FEDER PI17/00918/Cofinanciado FEDER; Acción Estratégica de Salud del Instituto de Salud Carlos III FIS Intrasalud (PI13/01136); Programa Grupos Emergentes, Cancer Genetics Unit, Instituto de Investigacion Biomedica Galicia Sur. Xerencia de Xestion Integrada de Vigo-SERGAS, Instituto de Salud Carlos III, Spain; Grant 10CSA012E, Consellería de Industria Programa Sectorial de Investigación Aplicada, PEME I + D e I + D Suma del Plan Gallego de Investigación, Desarrollo e Innovación Tecnológica de la Consellería de Industria de la Xunta de Galicia, Spain; Grant EC11-192. Fomento de la Investigación Clínica Independiente, Ministerio de Sanidad, Servicios Sociales e Igualdad, Spain; and Grant FEDER-Innterconecta. Ministerio de Economia y Competitividad, Xunta de Galicia, Spain. The BSUCH study was supported by the Dietmar-Hopp Foundation, the Helmholtz Society and the German Cancer Research Center (DKFZCBCS is funded by the Canadian Cancer Society (grant # 313404) and the Canadian Institutes of Health Research. CCGP is supported by funding from the University of Crete. The CECILE study was supported by Fondation de France, Institut National du Cancer (INCa), Ligue Nationale contre le Cancer, Agence Nationale de Sécurité Sanitaire, de l’Alimentation, de l’Environnement et du Travail (ANSES), Agence Nationale de la Recherche (ANR). The CGPS was supported by the Chief Physician Johan Boserup and Lise Boserup Fund, the Danish Medical Research Council, and Herlev and Gentofte Hospital. The American Cancer Society funds the creation, maintenance, and updating of the CPS-II cohort. The California Teachers Study (CTS) and the research reported in this publication were supported by the National Cancer Institute of the National Institutes of Health under award number U01-CA199277; P30-CA033572; P30-CA023100; UM1-CA164917; and R01-CA077398. The content is solely the responsibility of the authors and does not necessarily represent the oficial views of the National Cancer Institute or the National Institutes of Health. The collection of cancer incidence data used in the California Teachers Study was supported by the California Department of Public Health pursuant to California Health and Safety Code Section 103885; Centers for Disease Control and Prevention’s National Program of Cancer Registries, under cooperative agreement 5NU58DP006344; the National Cancer Institute’s Surveillance, Epidemiology and End Results Program under contract HHSN261201800032I awarded to the University of California, San Francisco, contract HHSN261201800015I awarded to the University of Southern California, and contract HHSN261201800009I awarded to the Public Health Institute. The opinions, findings, and conclusions expressed herein are those of the author(s) and do not necessarily reflect the oficial views of the State of California, Department of Public Health, the National Cancer Institute, the National Institutes of Health, the Centers for Disease Control and Prevention or their Contractors and Subcontractors, or the Regents of the University of California, or any of its programs. The University of Westminster curates the DietCompLyf database funded by Against Breast Cancer Registered Charity No. 1121258 and the NCRN. The coordination of EPIC is financially supported by the European Commission (DG-SANCO) and the International Agency for Research on Cancer. The national cohorts are supported by: Ligue Contre le Cancer, Institut Gustave Roussy, Mutuelle Générale de l’Education Nationale, Institut National de la Santé et de la Recherche Médicale (INSERM) (France); German Cancer Aid, German Cancer Research Center (DKFZ), Federal Ministry of Education and Research (BMBF) (Germany); the Hellenic Health Foundation, the Stavros Niarchos Foundation (Greece); Associazione Italiana per la Ricerca sul Cancro-AIRC-Italy and National Research Council (Italy); Dutch Ministry of Public Health, Welfare and Sports (VWS), Netherlands Cancer Registry (NKR), LK Research Funds, Dutch Prevention Funds, Dutch ZON (Zorg Onderzoek Nederland), World Cancer Research Fund (WCRF), Statistics Netherlands (The Netherlands); Health Research Fund (FIS), PI13/00061 to Granada, PI13/01162 to EPIC-Murcia, Regional Governments of Andalucía, Asturias, Basque Country, Murcia and Navarra, ISCIII RETIC (RD06/0020) (Spain); Cancer Research UK (14136 to EPIC-Norfolk; C570/A16491 and C8221/A19170 to EPIC-Oxford), Medical Research Council (1000143 to EPIC-Norfolk, MR/M012190/1 to EPIC-Oxford) (United Kingdom). The ESTHER study wasfunded by the Baden-Württemberg state Ministry of Science, Research and Arts (Stuttgart, Germany), the Federal Ministry of Research, Technology and Space (Berlin, Germany), the Federal Ministry of Education, Family Afairs, Senior Citizens, and Youth (Berlin, Germany), and the Saarland Ministry of Labour, Social Afairs, Women and Public Health. Additional cases were recruited in the context of the VERDI study, which was supported by a grant from the German Cancer Aid (Deutsche Krebshilfe). FHRISK and PROCAS are funded from NIHR grant PGfAR 0707-10031. DGE, AH and WGN are supported by the NIHR Manchester Biomedical Research Centre (IS-BRC-1215-20007). The GENICA was funded by the Federal Ministry of Education and Research (BMBF) Germany grants 01KW9975/5, 01KW9976/8, 01KW9977/0 and 01KW0114, the Robert Bosch Foundation, Stuttgart, Deutsches Krebsforschungszentrum (DKFZ), Heidelberg, the Institute for Prevention and Occupational Medicine of the German Social Accident Insurance, Institute of the Ruhr University Bochum (IPA), Bochum, as well as the Department of Internal Medicine, Johanniter GmbH Bonn, Johanniter Krankenhaus, Bonn, Germany. The GEPARSIXTO study was conducted by the German Breast Group GmbH. The GESBC was supported by the Deutsche Krebshilfe e. V. [70492] and the German Cancer Research Center (DKFZ). The HERPACC was supported by MEXT Kakenhi (No. 170150181 and 26253041) and by the Japan Society for the Promotion of Science (JSPS) KAKENHI Grant (No. 16H06277 and 22H04923 [CoBiA]) from the Ministry of Education, Science, Sports, Culture and Technology of Japan, by a Grant-in-Aid for the Third Term Comprehensive 10-Year Strategy for Cancer Control from Ministry Health, Labour and Welfare of Japan, by Health and Labour Sciences Research Grants for Research on Applying Health Technology from Ministry Health, Labour and Welfare of Japan, by National Cancer Center Research and Development Fund, and “Practical Research for Innovative Cancer Control (15ck0106177h0001 and 20ck0106553)” from Japan Agency for Medical Research and development, AMED, and Cancer Bio Bank Aichi. HKBCS study is supported by Dr. Ellen Li Charitable Foundation; Kerry Kuok Foundation; Health and Medical Research Fund (03143406); Asian Fund for Cancer Research, and Hong Kong Hereditary Breast Cancer Family Registry. The HMBCS was supported by the German Research Foundation (DFG Do761/15-1), a grant from the Friends of Hannover Medical School, and by the Rudolf Bartling Foundation. The HUBCS was supported by German Research Foundation (DFG Do761/15-1), a grant from the German Federal Ministry of Research and Education (RUS08/017), B.M. was supported by grant 17-44-020498, 17-29-06014 of the Russian Foundation for Basic Research, D.P. was supported by grant 18-29-09129 of the Russian Foundation for Basic Research, E.K was supported by the mega grant from the Government of Russian Federation (2020-220-08-2197), and the study was performed as part of the assignment of the Ministry of Science and Higher Education of the Russian Federation (№АААА-А16-116020350032-1). Financial support for KARBAC was provided through the regional agreement on medical training and clinical research (ALF) between Stockholm County Council and Karolinska Institutet, the Swedish Cancer Society, The Gustav V Jubilee foundation and Bert von Kantzows foundation. The KARMA study was supported by Märit and Hans Rausings Initiative Against Breast Cancer. The KBCP was financially supported by the special Government Funding (VTR) of Kuopio University Hospital grants, Cancer Fund of North Savo, the Finnish Cancer Organizations, and by the strategic funding of the University of Eastern Finland. The KOHBRA study was partially supported by a grant from the Korea Health Technology R&D Project through the Korea Health Industry Development Institute (KHIDI), and the National R&D Program for Cancer Control, Ministry of Health & Welfare, Republic of Korea (HI16C1127; 0720450; 1020350; 1420190). LMBC is supported by the ’Stichting tegen Kanker’. DL is supported by the FWO. The MABCS study is funded by the Research Centre for Genetic Engineering and Biotechnology “Georgi D. Efremov”, MASA. The MARIE study was supported by the Deutsche Krebshilfe e.V. [70-2892-BR I, 106332, 108253, 108419, 110826, 110828], the Hamburg Cancer Society, the German Cancer Research Center (DKFZ) and the Federal Ministry of Education and Research (BMBF) Germany [01KH0402]. MBCSG is supported by grants from the Italian Association for Cancer Research (AIRC) and the Italian Ministry of Health with Ricerca Corrente and 5×1000 Funds. The MCBCS was supported by the NIH grants R35CA253187, R01CA192393, R01CA116167, R01CA176785 a NIH Specialized Program of Research Excellence (SPORE) in Breast Cancer [P50CA116201], and the Breast Cancer Research Foundation. The Melbourne Collaborative Cohort Study (MCCS) cohort recruitment was funded by VicHealth and Cancer Council Victoria. The MCCS was further augmented by Australian National Health and Medical Research Council grants 209057, 396414 and 1074383 and by infrastructure provided by Cancer Council Victoria. Cases and their vital status were ascertained through the Victorian Cancer Registry. The MEC was supported by NIH grants CA63464, CA54281, CA098758, CA132839 and CA164973. The MISS study was supported by funding from ERC-2011-294576 Advanced grant, Swedish Cancer Society, Swedish Research Council, Local hospital funds, Fru Berta Kamprad Foundation, Gunnar Nilsson. The work of MTLGEBCS was supported by the Quebec Breast Cancer Foundation, the Canadian Institutes of Health Research for the “CIHR Team in Familial Risks of Breast Cancer” program – grant # CRN-87521 and the Ministry of Economic Development, Innovation and Export Trade – grant # PSR-SIIRI-701. The NBCS has received funding from the K.G. Jebsen Centre for Breast Cancer Research; the Research Council of Norway grant 193387/V50 (to V.N. Kristensen) and grant 193387/H10 (to V.N. Kristensen), and the Norwegian Cancer Society (to V.N. Kristensen). The Norwegian Health authorities 2014, 2018; the Norwegian Cancer Society, 2015, 2019: The genetic “Make up and metabolic profile of breast cancer patients; relation to clinical course and treatment response”. The NBHS was supported by NIH grant R01CA100374. Biological sample preparation was conducted the Survey and Biospecimen Shared Resource, which is supported by P30 CA68485. The Northern California Breast Cancer Family Registry (NC-BCFR) and Ontario Familial Breast Cancer Registry (OFBCR) were supported by grant U01CA164920 from the USA National Cancer Institute of the National Institutes of Health. The content of this manuscript does not necessarily reflect the views or policies of the National Cancer Institute or any of the collaborating centers in the Breast Cancer Family Registry (BCFR), nor does mention of trade names, commercial products, or organizations imply endorsement by the USA Government or the BCFR. The Carolina Breast Cancer Study (NCBCS) was funded by Komen Foundation, the National Cancer Institute (P50 CA058223, U54 CA156733, U01 CA179715), and the North Carolina University Cancer Research Fund. The NGOBCS was supported by the National Cancer Center Research and Development Fund (Japan). The NHS was supported by NIH grants P01 CA87969, UM1 CA186107, and U19 CA148065. The NHS2 was supported by NIH grants UM1 CA176726 and U19 CA148065. The ORIGO study was supported by the Dutch Cancer Society (RUL 1997-1505) and the Biobanking and Biomolecular Resources Research Infrastructure (BBMRI-NL CP16). The PBCS was funded by Intramural Research Funds of the National Cancer Institute, Department of Health and Human Services, USA. Genotyping for PLCO was supported by the Intramural Research Program of the National Institutes of Health, NCI, Division of Cancer Epidemiology and Genetics. The PLCO is supported by the Intramural Research Program of the Division of Cancer Epidemiology and Genetics and supported by contracts from the Division of Cancer Prevention, National Cancer Institute, National Institutes of Health. The POSH study is funded by Cancer Research UK (grants C1275/A11699, C1275/C22524, C1275/A19187, C1275/A15956 and Breast Cancer Campaign 2010PR62, 2013PR044. The RBCS was funded by the Dutch Cancer Society (DDHK 2004-3124, DDHK 2009-4318). The SBCGS was supported primarily by NIH grants R01CA64277, R01CA148667, UMCA182910, and R37CA70867. Biological sample preparation was conducted the Survey and Biospecimen Shared Resource, which is supported by P30 CA68485. The scientific development and funding of this project were, in part, supported by the Genetic Associations and Mechanisms in Oncology (GAME-ON) Network U19 CA148065. The SBCS was supported by Shefield Experimental Cancer Medicine Centre and Breast Cancer Now Tissue Bank. SEARCH is funded by Cancer Research UK [C490/A10124, C490/A16561] and supported by the UK National Institute for Health Research Biomedical Research Centre at the University of Cambridge. The University of Cambridge has received salary support for PDPP from the NHS in the East of England through the Clinical Academic Reserve. SEBCS was supported by the BRL (Basic Research Laboratory) program through the National Research Foundation of Korea funded by the Ministry of Education, Science and Technology (2012-0000347). SGBCC is funded by the Breast Cancer Screening and Prevention Programme, the National Research Foundation Singapore, NUS start-up Grant, National University Cancer Institute Singapore (NCIS) Centre Grant, Breast Cancer Prevention Programme, Asian Breast Cancer Research Fund and the NMRC Clinician Scientist Award (SI Category). Population-based controls were from the Multi-Ethnic Cohort (MEC) funded by grants from the Ministry of Health, Singapore, National University of Singapore and National University Health System, Singapore. The Sister Study (SISTER) is supported by the Intramural Research Program of the NIH, National Institute of Environmental Health Sciences (Z01-ES044005 and Z01-ES049033). The Two Sister Study (2SISTER) was supported by the Intramural Research Program of the NIH, National Institute of Environmental Health Sciences (Z01-ES044005 and Z01-ES102245), and, also by a grant from Susan G. Komen for the Cure, grant FAS0703856. The contributions of the NIH author(s) are considered Works of the United States Government. The findings and conclusions presented in this paper are those of the author(s) and do not necessarily reflect the views of the NIH or the U.S. Department of Health and Human Services. SKKDKFZS is supported by the DKFZ. The SMC is funded by the Swedish Cancer Foundation and the Swedish Research Council (VR 2017-00644) grant for the Swedish Infrastructure for Medical Population-based Life-course Environmental Research (SIMPLER). The SZBCS was supported by Grant PBZ_KBN_122/P05/2004 and the program of the Minister of Science and Higher Education under the name “Regional Initiative of Excellence” in 2019-2022 project number 002/RID/2018/19 amount of financing 12 000 000 PLN. The TNBCC was supported by: a Specialized Program of Research Excellence (SPORE) in Breast Cancer (CA116201), a grant from the Breast Cancer Research Foundation, a generous gift from the David F. and Margaret T. Grohne Family Foundation. The TWBCS is supported by the Taiwan Biobank project of the Institute of Biomedical Sciences, Academia Sinica, Taiwan. UBCS was supported by funding from National Cancer Institute (NCI) grant R01 CA163353 (to N.J. Camp) and the Women’s Cancer Center at the Huntsman Cancer Institute (HCI). Data collection for UBCS was supported by the Utah Population Database (UPDB) and Utah Cancer Registry (UCR). The UPDB is supported by HCI, the University of Utah, and NCI grant P30 CA2014. The UCR is additionally funded by the NCI’s SEER Program, HHSN261201800016I, and the US Center for Disease Control and Prevention’s National Program of Cancer Registries (NU58DP007131). The UCIBCS component of this research was supported by the NIH [CA58860, CA92044] and the Lon V Smith Foundation [LVS39420]. The UKBGS is funded by Breast Cancer Now and the Institute of Cancer Research (ICR), London. ICR acknowledges NHS funding to the NIHR Biomedical Research Centre. The UKOPS study was funded by The Eve Appeal (The Oak Foundation) and supported by the National Institute for Health Research University College London Hospitals Biomedical Research Centre. The USRT Study was funded by Intramural Research Funds of the National Cancer Institute, Department of Health and Human Services, USA. .

## Dava Availability

The source code of PRANA is available at: https://github.com/hag007/PRANA

Data are available upon reasonable request. The Breast Cancer Association Consortium (BCAC) data are available upon request from Cambridge University (see the BCAC website: https://bcac.ccge.medschl.cam.ac.uk/bcacdata/).

The UK biobank data were used by permission from https://www.ukbiobank.ac.uk/

## Acknowledgments

We thank Todd Lencz for providing us with the leave-one-out version of the SCZ GWAS. This research has been conducted using the UK Biobank application no. 56885.

R.E. is a Faculty Fellow of the Edmond J. Safra Center for Bioinformatics, Tel Aviv University. This work was carried out in partial fulfillment of the requirements for the Ph.D. degree of H.L. at the Blavatnik School of Computer Science, Tel Aviv University.

## Acknowledgements

We thank all the individuals who took part in these studies and all the researchers, clinicians, technicians and administrative staf who have enabled this work to be carried out. ABCFS thank Maggie Angelakos, Judi Maskiell, Gillian Dite. ABCS thanks Marjanka Schmidt, Linde Braaf, Annegien Broeks, Sander Canisius, Sten Cornelissen, Renate de Groot, Ellen Engelhardt, Renske Keeman, Sabine Linn, Efraim Rosenberg, Emiel Rutgers, Joyce Sanders, Vincent Smit, Alexandra van den Broek, C. Ellen van der Schoot, Laura Van’t Veer, Senno Verhoef, Jelle Wesseling, Maria Escala-Garcia, Anna Morra, Hugo Horlings, Marleen Kok, Frans Hogervorst, the Blood bank Sanquin, The Netherlands. ABCTB Investigators: Christine Clarke, Deborah Marsh, Rodney Scott, Robert Baxter, Desmond Yip, Jane Carpenter, Alison Davis, Nirmala Pathmanathan, Peter Simpson, J. Dinny Graham, Mythily Sachchithananthan. Samples are made available to researchers on a non-exclusive basis. The ACP study wishes to thank the participants in the Thai Breast Cancer study. Special thanks also go to the Thai Ministry of Public Health (MOPH), doctors and nurses who helped with the data collection process. Finally, the study would like to thank Dr Prat Boonyawongviroj, the former Permanent Secretary of MOPH and Dr Pornthep Siriwanarungsan, the former Department Director-General of Disease Control who have supported the study throughout. BBCS thanks Eileen Williams, Elaine Ryder-Mills, Kara Sargus. BCEES thanks Allyson Thomson, Christobel Saunders, Jennifer Girschik, Jane Heyworth and Terry Boyle. The BREOGAN study would not have been possible without the contributions of the following: Manuela Gago-Dominguez, Jose Esteban Castelao, Angel Carracedo, Victor Muñoz Garzón, Alejandro Novo Domínguez, Maria Elena Martinez, Sara Miranda Ponte, Carmen Redondo Marey, Maite Peña Fernández, Manuel Enguix Castelo, Maria Torres, Manuel Calaza (BREOGAN), José Antúnez, Máximo Fraga and the staf of the Department of Pathology and Biobank of the University Hospital Complex of Santiago-CHUS, Instituto de Investigación Sanitaria de Santiago, IDIS, Xerencia de Xestion Integrada de Santiago-SERGAS; Joaquín González-Carreró and the staf of the Department of Pathology and Biobank of University Hospital Complex of Vigo, Instituto de Investigacion Biomedica Galicia Sur, SERGAS, Vigo, Spain. The BSUCH study acknowledges the Principal Investigator, Barbara Burwinkel, and thanks Peter Bugert, Medical Faculty Mannheim CBCS thanks Kristan Aronson, Rachel Murphy, study participants, co-investigators, collaborators and staf of the Canadian Breast Cancer Study, and project coordinators Agnes Lai and Celine Morissette. CCGP thanks Styliani Apostolaki, Anna Margiolaki, Georgios Nintos, Maria Perraki, Georgia Saloustrou, Georgia Sevastaki, Konstantinos Pompodakis. CGPS thanks staf and participants of the Copenhagen General Population Study. For the excellent technical assistance: Dorthe Uldall Andersen, Maria Birna Arnadottir, Anne Bank, Dorthe Kjeldgård Hansen. The Danish Cancer Biobank is acknowledged for providing infrastructure for the collection of blood samples for the cases. The Danish Breast Cancer Cooperative Group (DBCG) are acknowledged for their provision of clinical case data. Investigators from the CPS-II cohort thank the participants and Study Management Group for their invaluable contributions to this research. They also acknowledge the contribution to this study from central cancer registries supported through the Centers for Disease Control and Prevention National Program of Cancer Registries, as well as cancer registries supported by the National Cancer Institute Surveillance Epidemiology and End Results program. The authors would like to thank the California Teachers Study Steering Committee that is responsible for the formation and maintenance of the Study within which this research was conducted. A full list of California Teachers Study (CTS) team members is available at https://www.calteachersstudy.org/team. DIETCOMPLYF thanks the patients, nurses and clinical staf involved in the study. The DietCompLyf study was funded by the charity Against Breast Cancer (Registered Charity Number 1121258) and the NCRN. We thank the participants and the investigators of EPIC (European Prospective Investigation into Cancer and Nutrition). ESTHER thanksBernd Holleczek, Karin Engel, Martina Mohr, and Utz Benscheid. FHRISK and PROCAS thank NIHR for funding. The GENICA Network: Dr. Margarete Fischer-Bosch-Institute of Clinical Pharmacology, Stuttgart, and University of Tübingen, Germany [Hiltrud Brauch, RH, Wing-Yee Lo], Department of Internal Medicine, Johanniter GmbH Bonn, Johanniter Krankenhaus, Bonn, Germany [Yon-Dschun Ko, Christian Baisch], Institute of Pathology, University of Bonn, Germany [Hans-Peter Fischer], Molecular Genetics of Breast Cancer, Deutsches Krebsforschungszentrum (DKFZ), Heidelberg, Germany [UH], Institute for Prevention and Occupational Medicine of the German Social Accident Insurance, Institute of the Ruhr University Bochum (IPA), Bochum, Germany [TB, Beate Pesch, Sylvia Rabstein, Anne Lotz]; and Institute of Occupational Medicine and Maritime Medicine, University Medical Center Hamburg-Eppendorf, Germany [Volker Harth]. . HKBCS thanks Hong Kong Sanatorium and Hospital; Genetic Counsellors and Medical Staf of Breast Center of Tung Wah Hospital, Hong Kong; Cecilia Ho, Dr. Edmond Ma and Staf of Molecular Pathology Laboratory, Hong Kong Sanatorium and Hospital; Dr. Sze Keong Tey and laboratory staf of Department of Surgery, Division of Breast Surgery, The University of Hong Kong, Hong Kong. HMBCS thanks Peter Hillemanns, Hans Christiansen and Johann H. Karstens. HUBCS thanks Darya Prokofyeva and Shamil Gantsev. KBCP thanks Eija Myöhänen. We thank all investigators of the KOHBRA (Korean Hereditary Breast Cancer) Study. LMBC thanks Gilian Peuteman, Thomas Van Brussel, EvyVanderheyden and Kathleen Corthouts. MABCS thanks Milena Jakimovska (RCGEB “Georgi D. Efremov”), Snezhana Smichkoska, Emilija Lazarova, Marina Iljoska (University Clinic of Radiotherapy and Oncology), Katerina Kubelka-Sabit, Dzengis Jasar, Mitko Karadjozov (Adzibadem-Sistina Hospital), Andrej Arsovski and Liljana Stojanovska (Re-Medika Hospital) for their contributions and commitment to this study. MARIE thanks Petra Seibold, Nadia Obi, Sabine Behrens, Ursula Eilber and Muhabbet Celik. MBCSG (Milan Breast Cancer Study Group): Siranoush Manoukian, Bernard Peissel, Jacopo Azzollini, Bernardo Bonanni, Irene Feroce, Mariarosaria Calvello, Aliana Guerrieri Gonzaga, Monica Marabelli, Matilde Risti and the personnel of the Cogentech Cancer Genetic Test Laboratory. The MCCS was made possible by the contribution of many people, including the original investigators, the teams that recruited the participants and continue working on follow-up, and the many thousands of Melbourne residents who continue to participate in the study. The MISS study group acknowledges the former Principal Investigator, Professor Håkan Olsson. We thank the coordinators, the research staf and especially the MSKCC thanks Marina Corines, Lauren Jacobs. MTLGEBCS would like to thank Martine Tranchant (CHU de Québec – Université Laval Research Center), Marie-France Valois, Annie Turgeon and Lea Heguy (McGill University Health Center, Royal Victoria Hospital; McGill University) for DNA extraction, sample management and skilful technical assistance. J.S. is Chair holder of the Canada Research Chair in Oncogenetics. The following are NBCS Collaborators: Kristine K. Sahlberg (PhD), Anne-Lise Børresen-Dale (Prof. Em.), Lars Ottestad (MD), Rolf Kåresen (Prof. Em.) Dr. Ellen Schlichting (MD), Marit Muri Holmen (MD), Toril Sauer (MD), Vilde Haakensen (MD), Olav Engebråten (MD), Bjørn Naume (MD), Alexander Fosså (MD), Cecile E. Kiserud (MD), Kristin V. Reinertsen (MD), Åslaug Helland (MD), Margit Riis (MD), Jürgen Geisler (MD), OSBREAC and Grethe I. Grenaker Alnæs (MSc). NBHS and SBCGS thank study participants and research staf for their contributions and commitment to the studies. For NHS and NHS2 the study protocol was approved by the institutional review boards of the Brigham and Women’s Hospital and Harvard T.H. Chan School of Public Health, and those of participating registries as required. We would like to thank the participants and staf of the NHS and NHS2 for their valuable contributions as well as the following state cancer registries for their help: AL, AZ, AR, CA, CO, CT, DE, FL, GA, ID, IL, IN, IA, KY, LA, ME, MD, MA, MI, NE, NH, NJ, NY, NC, ND, OH, OK, OR, PA, RI, SC, TN, TX, VA, WA, WY. The authors assume full responsibility for analyses and interpretation of these data. The OFBCR thanks Gord Glendon, Teresa Selander and the Sinai Health Biospecimen Repository, Nayana Weerasooriya and Steve Gallinger. ORIGO thanks E. Krol-Warmerdam, and J. Blom for patient accrual, administering questionnaires, and managing clinical information. The LUMC survival data were retrieved from the Leiden hospital-based cancer registry system (ONCDOC) with the help of Dr. J. Molenaar. PBCS thanks Louise Brinton, Mark Sherman, Neonila Szeszenia-Dabrowska, Beata Peplonska, Witold Zatonski, Pei Chao, Michael Stagner. The ethical approval for the POSH study is MREC /00/6/69, UKCRN ID: 1137. We thank staf in the Experimental Cancer Medicine Centre (ECMC) supported Faculty of Medicine Tissue Bank and the Faculty of Medicine DNA Banking resource. The authors wish to acknowledge the roles of the Breast Cancer Now Tissue Bank in collecting and making available the samples and/or data, and the patients who have generously donated their tissues and shared their data to be used in the generation of this publication. PREFACE thanks Sonja Oeser and Silke Landrith. The RBCS thanks Jannet Blom, Saskia Pelders, Wendy J.C. Prager – van der Smissen, and the Erasmus MC Family Cancer Clinic. SBCS thanks Sue Higham, Helen Cramp, Dan Connley, Ian Brock, Sabapathy Balasubramanian and Malcolm W.R. Reed. We thank the SEARCH and EPIC teams. SGBCC thanks the participants and all research coordinators for their excellent help with recruitment, data and sample collection. SKKDKFZS thanks all study participants, clinicians, family doctors, researchers and technicians for their contributions and commitment to this study. We thank the SUCCESS Study teams in Munich, Duessldorf, Erlangen and Ulm. UBCS thanks all study participants as well as the ascertainment, laboratory, analytics and informatics teams at Huntsman Cancer Institute and Intermountain Healthcare for their important contributions to this study. UCIBCS thanks Irene Masunaka. UKBGS thanks Breast Cancer Now and the Institute of Cancer Research for support and funding of the Generations Study, and the study participants, study staf, and the doctors, nurses and other health care providers and health information sources who have contributed to the study. We acknowledge NHS funding to the Royal Marsden/ICR NIHR Biomedical Research Centre.

## Supplementary Figures

**Figure S1.**
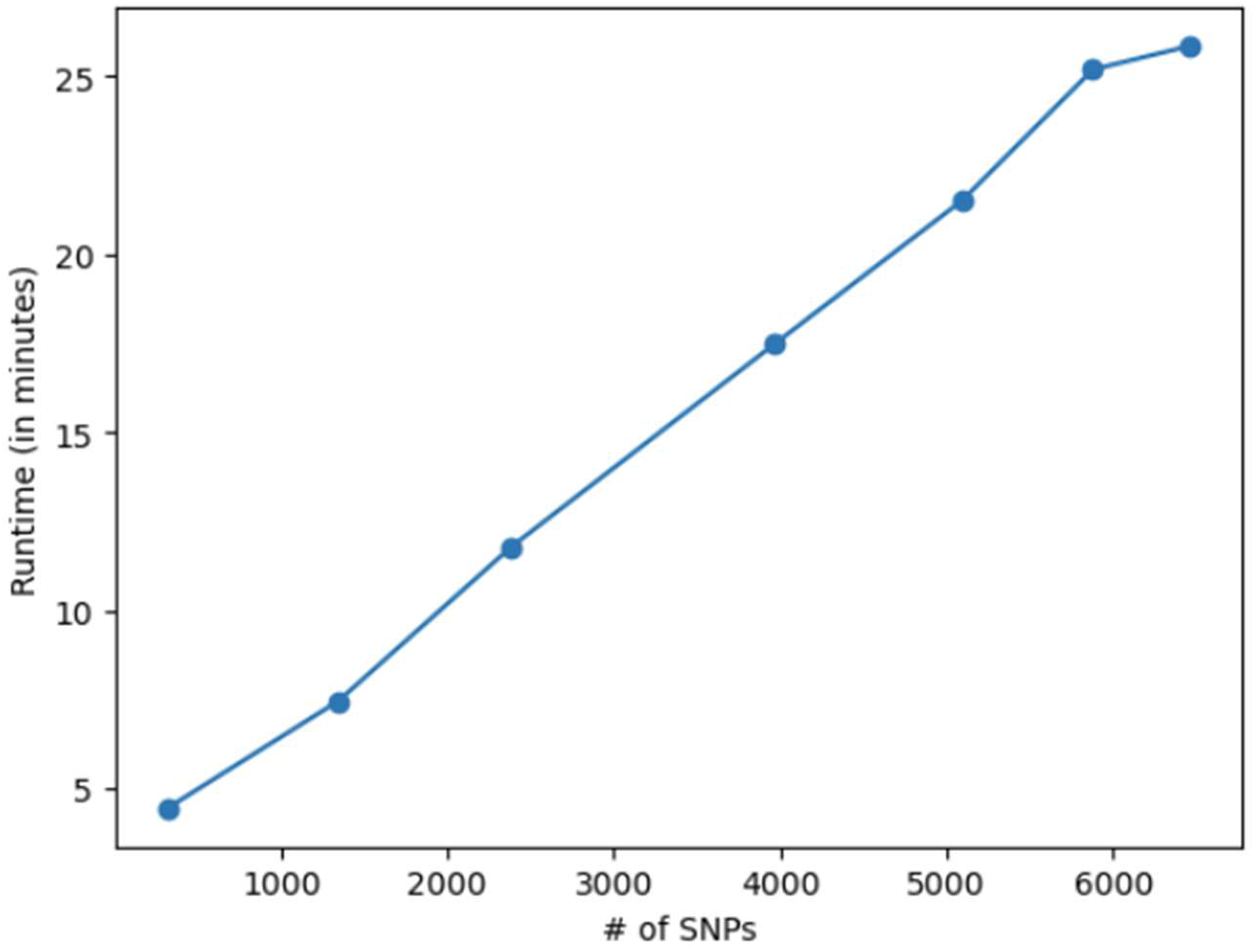
PRANA runtime as a function of the number of input SNPs. Runtimes were measured over 100 epochs on 8830 samples with 313 BC PRS. The number of SNPs was increased by incorporating into the model SNPs that are proximal to the 313 set and correlated with the original set.

## Notes

### Competing Interest Statement

The authors have declared no competing interest.

### Author Declarations

This study use only openly available data. The Breast Cancer Association Consortium (BCAC) data are available upon request from Cambridge University (see the BCAC website: https://bcac.ccge.medschl.cam.ac.uk/bcacdata/). AJ SCZ genotypes were derived by permission from dbgap (https://www.ncbi.nlm.nih.gov/gap/) using access ids phs000021.v3 phs000448.v1.p1 The UK biobank data were used by permission from https://www.ukbiobank.ac.uk/

